# Proteomic Profiling in Biracial Cohorts Implicates DC-SIGN as a Mediator of Genetic Risk in COVID-19

**DOI:** 10.1101/2020.06.09.20125690

**Authors:** Daniel H. Katz, Usman A. Tahir, Debby Ngo, Mark Benson, Alexander G. Bick, Akhil Pampana, Yan Gao, Michelle J. Keyes, Adolfo Correa, Sumita Sinha, Dongxiao Shen, Qiong Yang, Jeremy M. Robbins, Zsu-Zsu Chen, Daniel E. Cruz, Bennet Peterson, Pradeep Natarajan, Ramachandran S. Vasan, J. Gustav Smith, Thomas J. Wang, James G. Wilson, Robert E. Gerszten

**Author notes:** Correspondence: Robert E. Gerszten, MD, Division of Cardiovascular Medicine, Beth Israel Deaconess Medical Center, 185 Pilgrim Road, Baker 408, Boston, MA 02215.

## Abstract

COVID-19 is one of the most consequential pandemics in the last century, yet the biological mechanisms that confer disease risk are incompletely understood. Further, heterogeneity in disease outcomes is influenced by race, though the relative contributions of structural/social and genetic factors remain unclear.^1,2^ Very recent unpublished work has identified two genetic risk loci that confer greater risk for respiratory failure in COVID-19: the *ABO* locus and the 3p21.31 locus.^3^ To understand how these loci might confer risk and whether this differs by race, we utilized proteomic profiling and genetic information from three cohorts including black and white participants to identify proteins influenced by these loci. We observed that variants in the *ABO* locus are associated with levels of CD209/DC-SIGN, a known binding protein for SARS-CoV and other viruses,^4^ as well as multiple inflammatory and thrombotic proteins, while the 3p21.31 locus is associated with levels of CXCL16, a known inflammatory chemokine.^5^ Thus, integration of genetic information and proteomic profiling in biracial cohorts highlights putative mechanisms for genetic risk in COVID-19 disease.

## Introduction

SARS-CoV-2 infection displays a wide array of clinical manifestations and degrees of severity. While there is evidence that comorbidities, particularly cardiovascular and metabolic disease, are risk factors for disease severity and outcomes,^6^ the underlying biologic mechanisms that cause some to develop life threatening disease while others remain asymptomatic are not well understood.

The association recently observed between the *ABO* locus on chromosome 9 and susceptibility to respiratory failure in COVID-19 is consistent with earlier work showing an association between blood type and COVID-19 disease, though the mechanism(s) by which this locus might confer susceptibility to respiratory failure is unknown.^7,8^ The association with the 3p21.31 region observed in the same recent study was particularly novel, but also of unclear significance.^3^

Emerging proteomic technologies enable large-scale protein profiling in population-based studies. Leveraging available genetic data, investigators have identified the genetic architecture of the circulating proteome.^9–11^ Conversely, combing this information can identify proteomic signatures associated with specific loci or disease variants. Here we used measurements of 1,305 circulating proteins on the SOMAScan™ platform and genetic data from 4,859 participants in three large population-based studies: the Jackson Heart Study (JHS), a cohort of black participants, as well as meta-analyzed data from two white cohorts, the Framingham Heart Study (FHS) and the Malmö Diet and Cancer Study (MDCS). We tested for associations between genetic variants at the *ABO* and 3p21.31 loci and protein levels in the three cohorts to identify possible mediators of disease.

## Results

### Cohort Characteristics

Participants in the JHS with proteomics had an average age of 56 years and were 61% female. They had multiple comorbidities including hypertension (56% on treatment), diabetes mellitus (24%), and obesity (mean BMI 32). Baseline characteristics in FHS/MDCS have been reportedly previously.^9^ In brief, participants in FHS/MDCS were of similar age to JHS but with fewer females (49-53%), and lower prevalence of treated hypertension, obesity, and diabetes mellitus.

### pQTLs in the ABO locus

Table 1 shows the 56 proteins that associate with variants within 1MB of the transcription start site (TSS) of the *ABO* gene in either JHS or FHS/MDCS or both at a p-value < 5×10^−8^. Such variants are termed protein quantitative trait loci (pQTLs). Twenty-three proteins had significant genetic associations in both black and white subjects, while 15 were specific to JHS and 18 were specific to the FHS/MDCS. As might be expected given the *ABO* region’s known association with thrombosis,^12^ proteins associated with variants in this locus across all cohorts included ADAMTS13, von Willebrand Factor (vWF), Tie-1, Angiopoetin-1 receptor, VEGFR-2 and VEGFR-3. Inflammatory mediators, including P-selectin and E-selectin, Immunoglobulin superfamily containing leucine-rich repeat protein 2, and FAM3D (which has known cytokine activity) were also observed. Strikingly, CD209 antigen/DC-SIGN, which is the known binding site for multiple viruses including SARS-CoV, and a theorized binding site for SARS-CoV-2,^13^ showed a strong association in both white and black cohorts. The specific variant most strongly associated with levels differed between JHS and FHS/MDCS. To demonstrate the specificity of the aptamer for DC-SIGN protein, we separately identified 47 variants within the gene encoding DC-SIGN protein (on chromosome 19) that associated with DC-SIGN protein levels in JHS at genome wide significance (p < 5×10^−8^). Supplementary Table 1 summarizes data that similarly support aptamer specificity for all proteins described herein, using the presence of such variants at or near the gene coding for the target protein that also associate with measured protein levels (termed cis-pQTLs), mass spectrometry data, or immunoassay data.

**Table 1.**
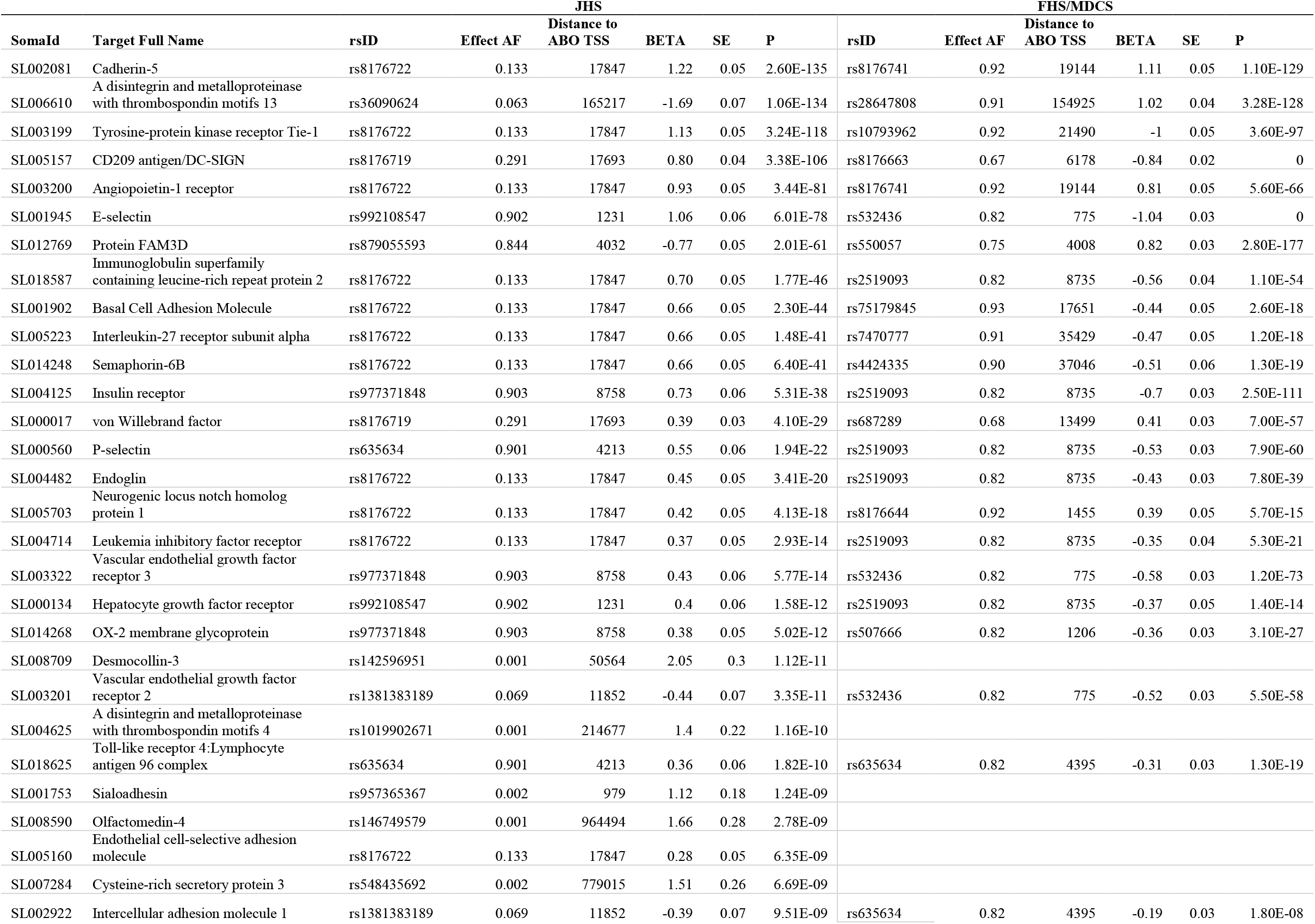
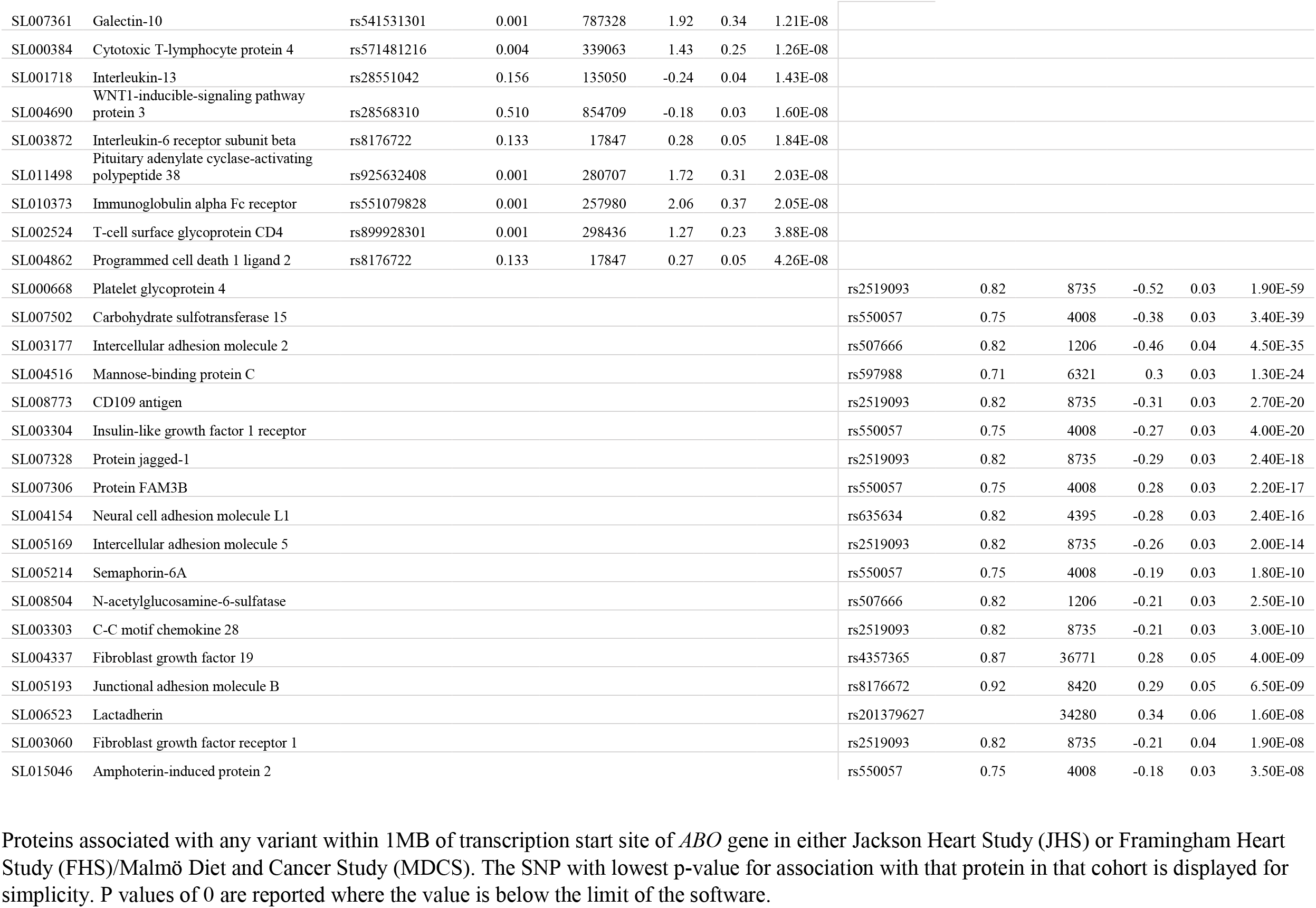
Proteins with pQTLs in the *ABO* Locus

Some protein associations were observed only in one racial group or the other. Among JHS participants, pQTLs in the *ABO* locus were associated with multiple inflammatory proteins including Cytotoxic T-lymphocyte protein 4, Interleukin-13, Interleukin-6 receptor subunit beta, Immunoglobulin alpha Fc receptor, T-cell surface glycoprotein CD4, and Programmed cell death 1 ligand 2. No pQTLs were identified for these proteins in FHS/MDCS. On the other hand, in FHS/MDCS but not JHS, variants in the *ABO* locus were associated with multiple adhesion molecules including Intercellular adhesion molecule 2, Neural cell adhesion molecule L1, Intercellular adhesion molecule 5, and Junctional adhesion molecule B.

### Proteins associated with rs657152

Given the above findings, we examined the specific variant identified by Ellinghaus et al., rs657152, which was the sentinel SNP at the *ABO* locus, having the strongest association with respiratory failure in COVID-19. Table 2 shows that this variant is a pQTL for a subset of the proteins associated with the larger locus described above. Notably, CD209 antigen/DC-SIGN, Basal Cell Adhesion Molecule, FAM3D, and vWF were all associated with this variant in both cohorts. For each of these four proteins, the effect allele that conferred higher risk for respiratory failure, A, was associated with higher measured levels of protein.

**Table 2.** Proteins Associated with rs657152

| SomaId | Target Full Name | JHS |  |  |  | FHS/MDCS |  |  |  |
| --- | --- | --- | --- | --- | --- | --- | --- | --- | --- |
|  |  | A Allele Frequency | BETA | SE | P | A Allele Frequency | BETA | SE | P |
| SL005157 | CD209 antigen/DC-SIGN | 0.42 | 0.50 | 0.03 | 1.77E-51 | 0.35 | 0.79 | 0.02 | 1.70E-271 |
| SL003199 | Tyrosine-protein kinase receptor Tie-1 | 0.42 | 0.45 | 0.03 | 1.19E-41 |  |  |  |  |
| SL002081 | Cadherin-5 | 0.42 | 0.36 | 0.03 | 7.38E-27 |  |  |  |  |
| SL001945 | E-selectin | 0.42 | -0.33 | 0.03 | 3.57E-22 |  |  |  |  |
| SL001902 | Basal Cell Adhesion Molecule | 0.42 | 0.29 | 0.03 | 4.73E-19 | 0.35 | 0.17 | 0.03 | 2.20E-10 |
| SL003200 | Angiopoietin-1 receptor | 0.42 | 0.28 | 0.03 | 2.56E-17 |  |  |  |  |
| SL012769 | Protein FAM3D | 0.42 | 0.25 | 0.03 | 6.84E-14 | 0.35 | 0.61 | 0.03 | 1.60E-108 |
| SL014248 | Semaphorin-6B | 0.42 | 0.23 | 0.03 | 6.62E-12 |  |  |  |  |
| SL000017 | von Willebrand factor | 0.42 | 0.22 | 0.03 | 8.20E-12 | 0.35 | 0.38 | 0.03 | 5.40E-49 |
| SL005223 | Interleukin-27 receptor subunit alpha | 0.42 | 0.18 | 0.03 | 4.29E-08 |  |  |  |  |
| SL004125 | Insulin receptor |  |  |  |  | 0.35 | -0.38 | 0.03 | 1.10E-48 |
| SL000560 | P-selectin |  |  |  |  | 0.35 | -0.33 | 0.03 | 1.10E-35 |
| SL000668 | Platelet glycoprotein 4 |  |  |  |  | 0.35 | -0.31 | 0.03 | 3.20E-34 |
| SL003322 | Vascular endothelial growth factor receptor 3 |  |  |  |  | 0.35 | -0.32 | 0.03 | 5.30E-33 |
| SL003201 | Vascular endothelial growth factor receptor 2 |  |  |  |  | 0.35 | -0.27 | 0.03 | 2.30E-25 |
| SL004516 | Mannose-binding protein C |  |  |  |  | 0.35 | 0.26 | 0.03 | 4.00E-23 |
| SL003177 | Intercellular adhesion molecule 2 |  |  |  |  | 0.35 | -0.28 | 0.03 | 3.40E-20 |
| SL007306 | Protein FAM3B |  |  |  |  | 0.35 | 0.23 | 0.03 | 1.90E-14 |
| SL014268 | OX-2 membrane glycoprotein |  |  |  |  | 0.35 | -0.19 | 0.03 | 2.90E-13 |
| SL018587 | Immunoglobulin superfamily containing leucine-rich repeat protein 2 |  |  |  |  | 0.35 | -0.19 | 0.03 | 2.70E-10 |
| SL004482 | Endoglin |  |  |  |  | 0.35 | -0.16 | 0.03 | 1.50E-09 |
| SL003303 | C-C motif chemokine 28 |  |  |  |  | 0.35 | -0.15 | 0.03 | 7.90E-09 |
| SL008504 | N-acetylglucosamine-6-sulfatase |  |  |  |  | 0.35 | -0.15 | 0.03 | 1.50E-08 |
| SL003304 | Insulin-like growth factor 1 receptor |  |  |  |  | 0.35 | -0.15 | 0.03 | 2.60E-08 |
| SL004154 | Neural cell adhesion molecule L1 |  |  |  |  | 0.35 | -0.15 | 0.03 | 2.70E-08 |
| SL000134 | Hepatocyte growth factor receptor |  |  |  |  | 0.35 | -0.2 | 0.04 | 3.10E-08 |
| SL005169 | Intercellular adhesion molecule 5 |  |  |  |  | 0.35 | -0.15 | 0.03 | 3.30E-08 |

Proteins with association with rs657152 in either Jackson Heart Study (JHS) or Framingham Heart Study (FHS)/Malmö Diet and Cancer Study (MDCS) meta-analysis. All Beta estimates are for the presence of the A allele.

### pQTLs in the chr3:45800446-46135604 locus

Given the multiple genes spanned by the other susceptibility locus, on chromosome 3, we similarly looked for pQTLs in this locus. Table 3 shows the proteins associated with variants in this region. Two proteins were found to have pQTLs in this locus across all cohorts: C-X-C motif chemokine 16 (CXCL16) and Teratocarcinoma growth factor 1. The *TDGF1* gene is near this locus making this a cis-pQTL. Further, while not a true cis relationship, CXCL16 is the ligand for CXCR6, whose gene is within this locus. No proteins were significantly associated with the specific variant identified by Ellinghaus et al., rs11385942.

**Table 3.** Proteins with pQTLs in the *chr3:45800446-46135604* Locus of hg38

| JHS |  |  |  |  |  |  | FHS/MDCS |  |  |  |  |
| --- | --- | --- | --- | --- | --- | --- | --- | --- | --- | --- | --- |
| SomaId | Target Full Name | rsID | Effect AF | BETA | SE | P | rsID | Effect AF | BETA | SE | P |
| SL004016 | C-X-C motif chemokine 16 | rs2234355 | 0.44 | 0.5 | 0.03 | 8.78E-50 | rs4396933 | 0.69 | 0.18 | 0.03 | 6.50E-09 |
| SL005155 | Teratocarcinoma-derived growth factor 1 | rs1546079 | 0.11 | 0.47 | 0.05 | 2.20E-18 | rs13434029 | 0.67 | -0.28 | 0.03 | 5.49E-19 |
| SL000076 | Cyclin-dependent kinase inhibitor 1B | rs9870445 | 0.86 | 0.31 | 0.04 | 5.49E-12 |  |  |  |  |  |
| SL004739 | Inter-alpha-trypsin inhibitor heavy chain H4 | rs1860264 | 0.83 | -0.25 | 0.04 | 1.90E-08 |  |  |  |  |  |
| SL007361 | Galectin-10 | rs373364694 | 0.001 | 1.89 | 0.34 | 2.01E-08 |  |  |  |  |  |
| SL004356 | C-C motif chemokine 4-like |  |  |  |  |  | rs1500004 | 0.88 | 0.55 | 0.05 | 5.40E-28 |

Proteins associated with any variant within *chr3:45800446-46135604* of hg38 in either Jackson Heart Study (JHS) or Framingham Heart Study (FHS)/Malmö Diet and Cancer Study (MDCS) meta-analysis. The SNP with lowest p-value for association with that protein in that cohort is displayed for simplicity. As GWAS in FHS/MDCS was performed using hg37, the locus has been translated to *chr3:45841939-46177097*.

Finally, we examined whether circulating levels of CXCL16 or DC-SIGN are associated with known risk factors for COVID-19 in JHS using unadjusted associations. While CXCL16 showed modest associations with age, sex, BMI, smoking, and renal function, DC-SIGN only showed a weak association with coronary disease (Table 4).

**Table 4.** Associations between clinical risk factors and protein levels of DC-SIGN and CXCL16 in the Jackson Heart Study

| Variable | *DC-SIGN* |  | *CXCL16* |  |
| --- | --- | --- | --- | --- |
|  | Coefficient | P Value | Coefficient | P Value |
| Age (y) | -0.03 | 0.2 | 0.08 | 0.0002* |
| Gender (male) | -0.06 | 0.14 | -0.29 | 2.00E-11* |
| Body Mass Index (m<sup>2</sup>/kg) | -0.03 | 0.17 | -0.19 | <2E-16* |
| Systolic Blood pressure (mmHg) | -0.003 | 0.88 | 0.26 | 0.24 |
| HbA1c(%) | -0.01 | 0.57 | 0.02 | 0.25 |
| Estimated glomerular filtration rate (mL/min/1.73m<sup>2</sup>) | 0.02 | 0.27 | -0.16 | 2.30E-14* |
| Total Cholesterol (mg/dL) | 0.03 | 0.12 | -0.02 | 0.45 |
| Coronary Heart Disease History | 0.09 | 0.02* | 0.09 | 0.70 |
| Current Smoker | 0.01 | 0.89 | 0.14 | 0.02* |

Unadjusted associations shown, using Pearson correlations for continuous variables and logistic regression for dichotomous variables. All protein levels were log-transformed and scaled by batch to normalize the data and reduce batch effects. \*Highlights p-values <0.05

## Discussion

The COVID-19 pandemic is a continually evolving public health crisis, and the biological mechanisms that confer the heterogeneous outcomes of infection remain unclear. Given the recently identified COVID-19 risk loci, our data identify several possible pathways by which these loci might confer risk in COVID-19.

The *ABO* locus is highly pleiotropic in our pQTL data, being associated with the levels of 56 proteins across the black and white cohorts. This likely reflects, in part, its role as a glycosyltransferase, altering the overall structure of multiple glycoproteins. An association between ABO blood group and disease is emerging in COVID-19. At least one published study observed a higher proportion of type A blood among individuals hospitalized with COVID-19,^14^ and unpublished data from China and the United States suggests that blood group A is associated with increased risk of acquiring COVID-19. In China, an increased proportion of Type A blood was observed among those with COVID-19 as compared with local controls.^8^ Zietz and Tatonetti found similar results among patients from New York Presbyterian Hospital, and meta-analyzed with the data from China to confirm the association.^7^ Most recently, the *ABO* locus was shown to be a risk locus for COVID-19 severity; in a meta-analysis of 1,610 cases of COVID-19 and respiratory failure and 2,205 COVID-19 cases without respiratory failure across a population of patients from Spain and Italy, Ellinghaus et al. observed multiple variants in the *ABO* gene locus that conferred increased risk.^3^ They, and others, have speculated about the possible ways in which ABO might play a role in COVID-19. The thrombotic risk associated with this locus is one plausible element, and indeed we observe the well-known association of this locus with vWF levels.^12,15^ Others suggest that blood type associates with ACE2 levels.^16^ Still others hypothesized that circulating anti-A antibodies in individuals with Type O or B blood might also confer some level of immunity to COVID-19.^14,17^ Our data, however, strongly suggest that *ABO* influences CD209 antigen/DC-SIGN, which is a known binding site for SARS-CoV.^4^ DC-SIGN is expressed by dendritic cells, and it has been shown that the SARS-CoV spike protein utilizes this protein for cell entry, as do HIV and dengue virus.^4^ Interestingly, DC-SIGN is thought to increase with age, and pre-print data suggest that it is also increased in smokers, associating it with two risk factors for COVID-19,^18^ though we did not observe these associations in our JHS data. Additionally, we did not observe an association with ACE2 levels measured on the SOMAScan™ platform. The association between *ABO* and DC-SIGN is consistent with data from other GWAS studies of the human proteome,^10,11^ and we now show the association in a black population. The frequency of the risk allele is slightly higher in JHS, one potential (though likely modest) reason for racial differences in COVID-19. Taken together, these data suggest that the *ABO* locus may confer its risk in part by modulating DC-SIGN, a putative binding site for SARS-CoV-2 cell entry.

Our data further suggest that the *ABO* locus may influence disease through pleiotropic effects, which may differ between black and white individuals. We show that *ABO* variants are associated with proteins involved in endothelial function and thrombosis, important complicating factors in COVID-19.^19^ However, there were key differences between the cohorts. In JHS, levels of multiple inflammatory proteins were associated with *ABO* variants, whereas in FHS/MDCS, *ABO* variation was associated with proteins involved in cellular adhesion. These differences may mediate increased risk, perhaps for inflammatory complications of COVID-19, such as cytokine storm.^20^

We also observed interesting associations at the other risk locus identified by Ellinghaus et al., at which the sentinel effect allele conferred higher risk in their analysis, compared to *ABO*.^3^ The validity of the 3p21.31 locus is noted by them to be supported by the Covid-19 Host Genetics Consortium data, which also showed increased COVID-19 risk, albeit at a lower level of significance.^21^ In our analysis, two key proteins emerged, CXCL16 and Teratocarcinoma growth factor 1. TDGF-1 is a signaling protein, and has an unclear relationship to infection or propagation of respiratory disease. CXCL16, on the other hand, is the chemokine ligand for the CXCR6 receptor, whose coding gene is at the chromosome 3 locus of interest. This makes the *CXCR6* gene a strong candidate for further investigation among the large number of genes at this locus. Indeed CXCL16/CXCR6 has been implicated in LPS-induced acute lung injury and alveolar inflammation previously.^5,22^

### Limitations

Our study has several important limitations. We have not studied the relationship between these proteins and genes in COVID-19 cases, thus confirming their role in pathogenesis requires further study. While the proteomic profiling discussed here is extensive, it does not cover the entire proteome, and important protein associations may be missed as a result. Further, while changes in aptamer binding are typically reflective of protein levels, it may be the case that ABO-mediated glycosylation alters aptamer binding, thus changing the measurement of the protein without a true change in protein levels. Nonetheless, our data demonstrate that ABO affects the identified proteins, motivating further investigation. Additionally, in the case of DC-SIGN (and CXCL16), we and others have found that variants at the cognate gene are associated with levels of the protein as measured by the aptamer (Supplementary Table 1). Finally, we acknowledge we do not observe an association between the lead risk SNP at 3p21.31 identified by Ellinghaus et al. and CXCL16. Formal colocalization in larger cohorts will be useful in clarifying the gene-protein relationships.

## Conclusions

We show here extensive proteomic profiling of genetic variation proposed to confer risk in COVID-19. We have shown in black and white cohorts that the *ABO* locus is associated with DC-SIGN, a putative binding site for SARS-CoV-2, suggesting ABO-mediated alteration of DC-SIGN may play a key role in disease pathogenesis. We further identify the CXCL16/CXCR6 pair as another potential disease mediator. Further study, specifically in COVID-19 infected patients, is needed to confirm these findings and determine whether levels or other modifications of these proteins could alter disease processes.

## Methods

### Study Approval

The human study protocols were approved by the Institutional Review Boards of Beth Israel Deaconess Medical Center, Boston University Medical Center, Lund University, and University of Mississippi Medical Center, and all participants provided written informed consent.

### Proteomic samples

The Jackson Heart Study is a community-based longitudinal cohort study begun in 2000 of 5306 self-identified African Americans from the Jackson, Mississippi metropolitan statistical area, the design of which is previously described.^23^ Baseline characteristics were assessed at Visit 1 between 2000 and 2004. Included in the present study are 1813 individuals with proteomic profiling and whole genome sequencing. Resting blood pressure was measured while sitting by recording two measurements with a Hawksley random zero sphygmomanometer using one of four cuff sizes selected by measured arm circumference. Glomerular filtration rate was estimated using the CKD-EPI equation.^24^ Prevalent coronary heart disease (CHD) at Visit 1 was determined as a composite of patient reported angina, patient reported myocardial infarction, and evidence of previous myocardial infarction on ECG. JHS plasma samples were collected at Visit 1 in EDTA tubes maintained in −70°C freezers.^25^ Proteomic measurements were performed using SOMAscan™, a single-stranded DNA aptamer-based proteomics platform, which contained 1,305 aptamers.^26^ Samples were run in three separate batches for cost efficiency. Batch 1 was run as a nested case-control study of incident coronary disease, excluding those with prevalent coronary heart disease at Visit 1. Batches 2 and 3 were a randomly selected sample of the remaining JHS participants. Each batch was divided into several plates containing a subset of the samples. The FHS Offspring study has been previously described.^27,28^ Included in the present study are 1625 individuals with proteomic profiling and genotyping performed at Visit 5. Proteomic profiling in FHS was also performed on the SOMAscan™ platform. In FHS, plasma samples were collected in citrate-treated tubes, which were then centrifuged within 15 minutes at 2000 g for 10 minutes and the supernatant plasma was aliquoted and stored at −80°C without freeze thaw cycles until assayed. This was completed in 2 different batches. In batch one, 1129 proteins (1.1k) were profiled in 695 individuals. As a result of platform enhancements that occurred in the interval between the first and second set of samples being run, batch 2 included an expanded panel of 1305 proteins (1.3k), which was assayed in 930 participants. The MDCS is a Swedish population-based, prospective, observational cohort recruited between 1991 and 1996.^29^ Included in the present study are 1421 individuals with proteomics and genotyping. Proteomic profiling in MDCS was also performed on the SOMAscan™ 1.3k platform as above, except samples were collected in EDTA-treated tubes. All assays in all cohorts were performed using SOMAscan™ reagents according to the manufacturer’s detailed protocol.^30^

### Genotyping and Imputation

Whole genome sequencing (WGS) in JHS has been described previously.^31^ JHS participants underwent 30× WGS through the Trans-Omics for Precision Medicine (TOPMed) project at the Northwest Genome Center at University of Washington; genotype calling was performed by the Informatics Resource Center at the University of Michigan.

Genome-wide genotyping methods for the FHS have been described previously.^32^ Briefly, genotyping was conducted using the Affymetrix 500K mapping array and the Affymetrix 50K gene-focused MIP supplemental array. Genotypes were called using Chiamo (http://www.stats.ox.ac.uk/~marchini/software/gwas/chiamo.html). We used the 1000 Genomes Phase I version 3 (August 2012) reference panel to perform imputation using a hidden Markov model implemented in MaCH (version 1.0.16)^33^ for all SNPs passing the following criteria: call rate ≥ 97%, pHWE ≥ 1 × 10^−6^, Mishap P ≥ 1 × 10−9, Mendel errors ≤ 100, and MAF ≥ 1%.

In the MDCS, genotyping was conducted using the Illumina Omni Express Exome BeadChip kit. Genotypes were called using Illumina GenomeStudio and imputation performed to the same 1000 Genomes version as for FHS using IMPUTE (v2) for SNPs passing the following criteria: call rate ≥ 95%, pHWE ≥ 1 × 10^−6^, minor allele frequency ≥ 0.01.

### Statistical analysis

In JHS, because proteomic data varied by batch, measurements were first standardized to a set of control samples that were part of each plate. Because of the non-normal distribution of the resulting protein levels, age, sex, and batch adjusted residuals were generated and inverse normalized. The association between these values and genetic variants was tested using linear mixed effects models adjusted for age, sex and the genetic relationship matrix to adjust for relatedness using the fastGWA model implemented in the GCTA software package.^34^ Variants with a minor allele count less than 5 were excluded.

In FHS and MDCS, inverse normalized transformed values of protein levels were also used. The association of genetic variants and protein levels were tested using linear mixed effects models to accommodate pedigree structure under an additive genetic model, adjusted for age and sex. Genome-wide association analyses were performed using the R GWAF package.^35^

Association analyses were limited to within 1MB of the transcription start site (TSS) of the ABO locus on chromosome 9 as well as the locus on chromosome 3 identified by Ellinghaus et al.: chr3:45800446-46135604 in Build hg38. Since FHS loci are identified by Build hg37 location, the region was shifted using the sentinel SNP rs11385942 as the reference point to chr3:45841939-46177097 in Build hg37. As we had hypothesis-driven genomic loci of interest, statistical significance for protein quantitative trait loci (pQTLs) was kept at a level of 5×10^−8^ despite multiple comparisons.

To determine associations between clinical variables and DC-SIGN or CXCL16, we used Pearson correlations for continuous variables and logistic regression for dichotomous variables. All protein levels were log-transformed and scaled by batch to normalize the data and reduce batch effects.

## Data Availability

Genetic and cohort data for JHS and FHS are available through dbGaP and TOPMed, with approvals from the respective cohorts required. Recently completed proteomic data in all cohorts is not publicly available at this time but is being prepared upload to dbGaP. For non-publicly available data inquiries, please contact the corresponding authors.

https://www.nhlbiwgs.org/

https://dbgap.ncbi.nlm.nih.gov/

https://framinghamheartstudy.org/

https://www.jacksonheartstudy.org/

## Acknowledgements

The Jackson Heart Study (JHS) is supported and conducted in collaboration with Jackson State University (HHSN268201800013I), Tougaloo College (HHSN268201800014I), the Mississippi State Department of Health (HHSN268201800015I/HHSN26800001) and the University of Mississippi Medical Center (HHSN268201800010I, HHSN268201800011I and HHSN268201800012I) contracts from the National Heart, Lung, and Blood Institute (NHLBI) and the National Institute for Minority Health and Health Disparities (NIMHD). The Framingham Heart Study (FHS) acknowledges the support of contracts NO1-HC-25195, HHSN268201500001I and 75N92019D00031 from the National Heart, Lung and Blood Institute and grant supplement R01 HL092577-06S1 for this research. Dr. Katz is supported by NHLBI T32 postdoctoral training grant (T32HL007374-40). Dr. Tahir is supported by the Ruth L. Kirchstein post-doctoral individual National Research Award (F32HL150992). Dr. Cruz is supported by the KL2/Catalyst Medical Research Investigator Training award from Harvard Catalyst (NIH/NCATS Award TR002542). Dr. Robbins is supported by the John S. LaDue Memorial Fellowship in Cardiology at Harvard Medical School. Dr. Benson is supported by NHLBI K08HL145095 award. J. Gustav Smith was supported by grants from the Swedish Heart-Lung Foundation (2016-0134, 2016-0315 and 2019-0526), the Swedish Research Council (2017-02554), the European Research Council (ERC-STG-2015-679242), the Crafoord Foundation, Skåne University Hospital, the Scania county, governmental funding of clinical research within the Swedish National Health Service, a generous donation from the Knut and Alice Wallenberg foundation to the Wallenberg Center for Molecular Medicine in Lund, and funding from the Swedish Research Council (Linnaeus grant Dnr 349-2006-237, Strategic Research Area Exodiab Dnr 2009-1039) and Swedish Foundation for Strategic Research (Dnr IRC15-0067) to the Lund University Diabetes Center. Drs. Gerszten, Wang and Wilson are supported by NIH R01 DK081572. Drs. Gerszten, Wang, and Ramachandran are supported by NIH R01 HL132320.

Molecular data for the Trans-Omics in Precision Medicine (TOPMed) program was supported by the National Heart, Lung and Blood Institute (NHLBI). Genome sequencing for “NHLBI TOPMed: The Jackson Heart Study” (phs000964.v1.p1) was performed at the Northwest Genomics Center (HHSN268201100037C). Core support including centralized genomic read mapping and genotype calling, along with variant quality metrics and filtering were provided by the TOPMed Informatics Research Center (3R01HL-117626-02S1; contract HHSN268201800002I). Core support including phenotype harmonization, data management, sample-identity QC, and general program coordination were provided by the TOPMed Data Coordinating Center (R01HL-120393; U01HL-120393; contract HHSN268201800001I). We gratefully acknowledge the studies and participants who provided biological samples and data for TOPMed.

The authors wish to thank the staffs and participants of the JHS.

## Disclaimer

*The views expressed in this manuscript are those of the authors and do not necessarily represent the views of the National Heart, Lung, and Blood Institute; the National Institutes of Health; or the U*.*S. Department of Health and Human Services*

---

**Supplementary Table 1.**
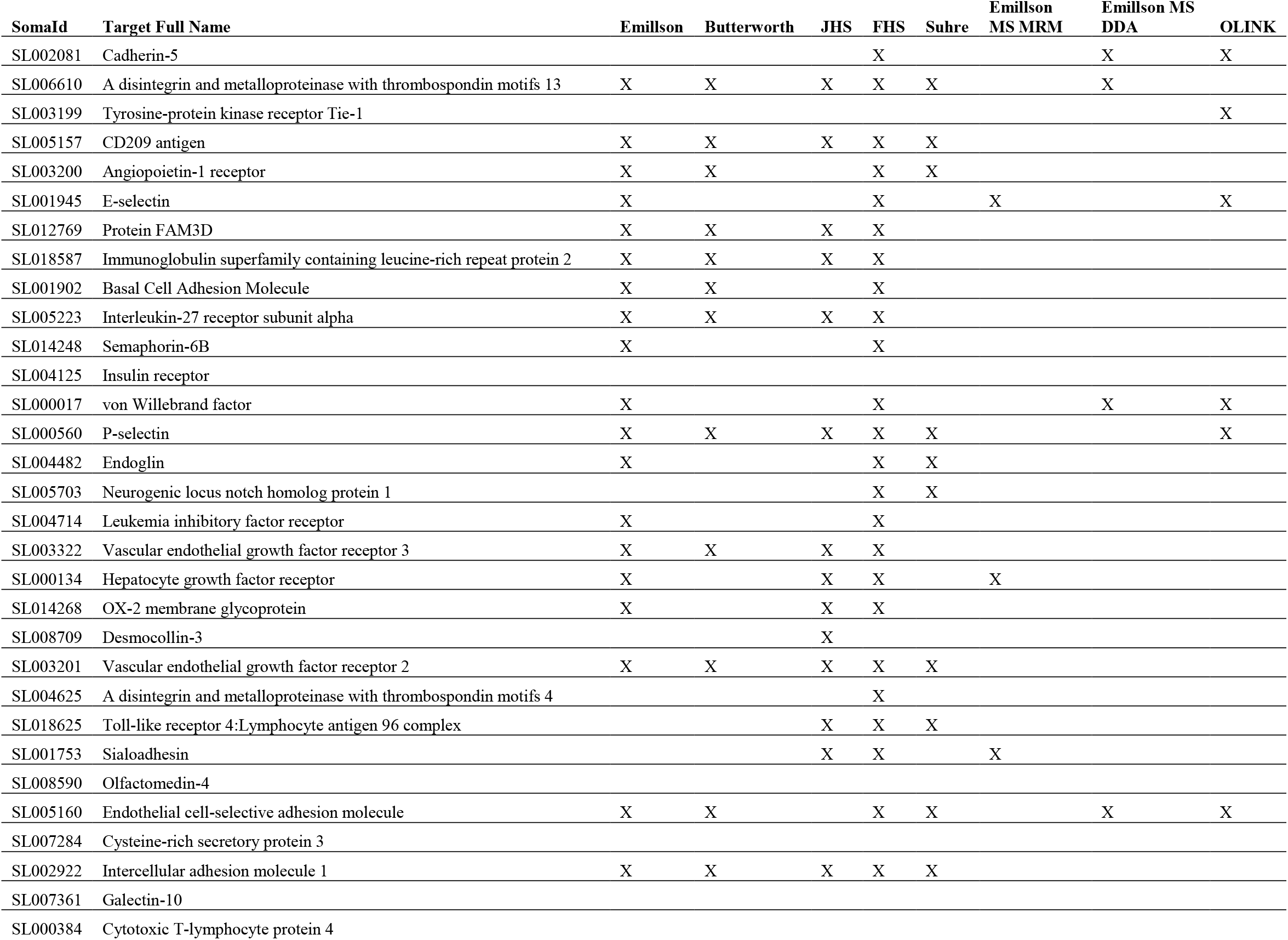
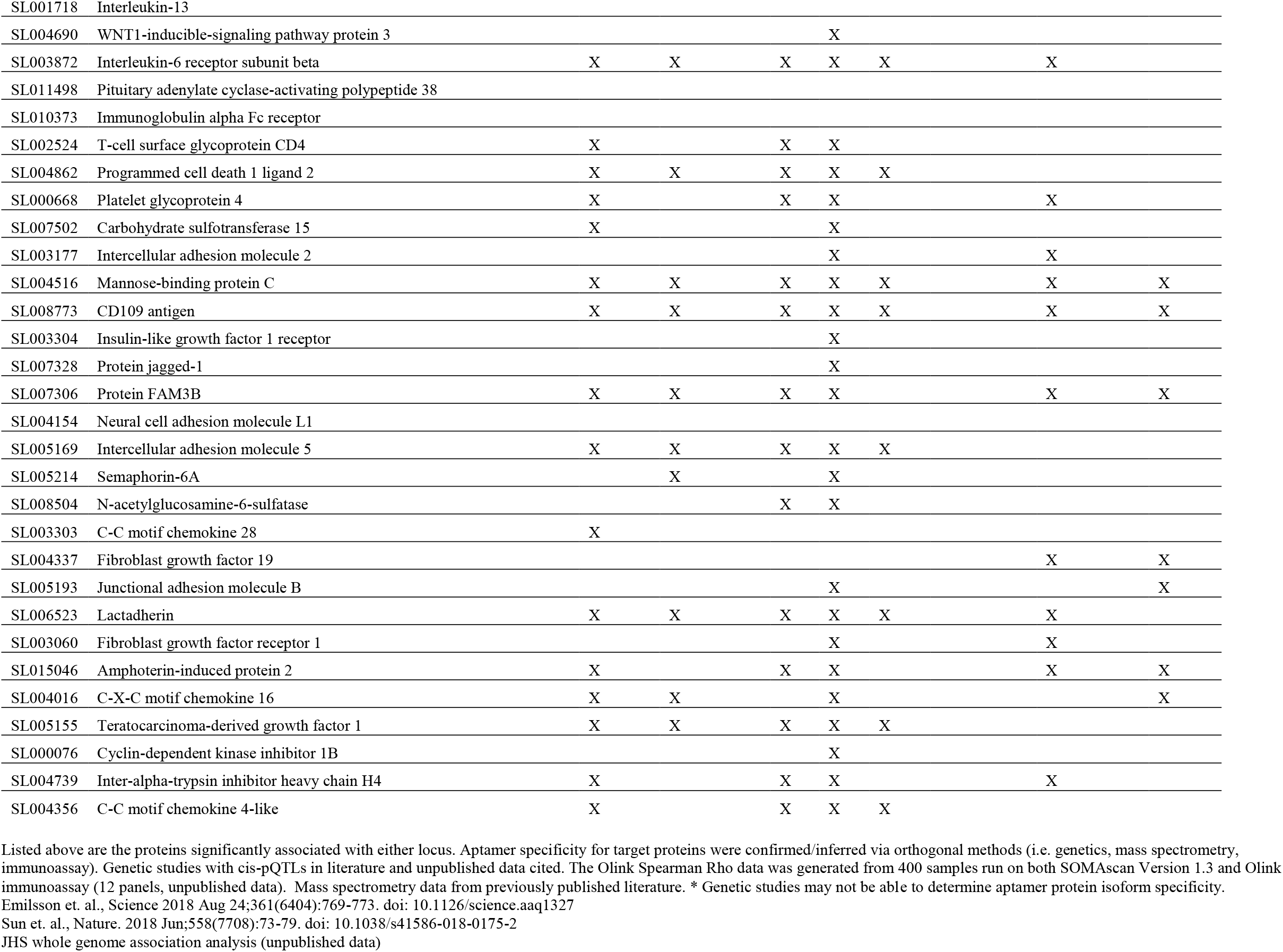
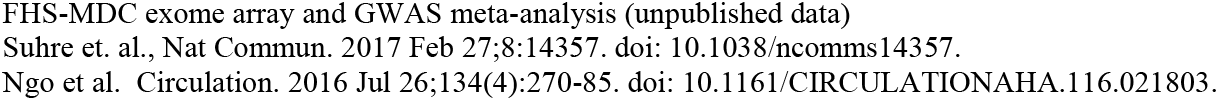
Data Sources Corroborating Aptamer Specificity

